# Assessing the Impact of Haulage drivers in Uganda’s COVID-19 Delta Wave

**DOI:** 10.1101/2024.09.10.24313441

**Authors:** Adrian Muwonge, Paul R Bessell, Mark Barend de Clare Bronsvoort, Ibrahim Mugerwa, Erisa Mwaka, Emmanuel Ssebaggala, Bryan Aidan Wee, Aggelos Kiayias, Christine Mbabazi Mpyangu, Moses Lutakome Joloba

## Abstract

**Background:** Haulage truck drivers can quickly connect distant communities, with risks of potential disease introduction. However, interventions to limit such risk must balance public health protection, economic continuity, and individual rights. Here distinguishing between their role in disease introduction and its onward spread is crucial for achieving this balance.

**Methods:** To investigate the role of haulage during the Delta wave of COVID-19 in Uganda. We fit a susceptible-infectious-recovered (SIR) model to the 625,422 records in the national surveillance dataset to assess the notion of a “core-risk group” by examining the incidence and impact of haulage-targeted interventions in border districts associated with heavy haulage traffic compared to the districts in the central region of Uganda.

**Results:** Although haulage drivers accounted for only 0.036% of the cases, the border districts associated with them registered 12.02% more cases than inland districts, suggesting a role in disease introduction. This risk was particularly higher in Tororo, compared to Amuru and Kyotera, which border Kenya, South Sudan, and Tanzania, respectively. Some interventions even increased the risk in Tororo by as much as 6%. However, in general, the haulage targeted interventions reduced the case load in border districts but registered limited impact on inland districts. This suggests a limited role in secondary within country spread. We note that combining such interventions with vaccination achieved greater reduction in case load.

**Conclusions:** Our findings suggest that truck drivers were a core risk group, though this risk was transient and in some cases exacerbated by some interventions. Pandemic preparedness strategies should characterize risks posed by core groups to ensure interventions balance public safety with individual rights in key sectors like supply chains.

## Introduction

To develop robust public health preparedness strategies, lessons must be drawn from the recent COVID-19 pandemic. These lessons are crucial for future strategies to: a) efficiently allocate resources, b) rapidly integrate data for informed decision-making, c) minimize the impact of population-wide movement restrictions and most importantly, d) improve risk attribution for tailored pandemic responses [1–4]. Countries like Uganda have developed response strategies due to frequent disease outbreaks of Ebola, Congo-Crimean Haemorrhagic Fever, and Marburg Virus Disease [5]. However, these strategies proved inadequate when faced with the scale and speed of the COVID-19 pandemic. COVID-19 especially tested the ability to balance public health, economic benefit, and individual rights during population-wide movement restrictions. Only essential workers operating critical systems such as healthcare, national security and supply chains, were exempted from such restrictions. However, disproportionately greater scrutiny was directed towards some supply chain workers such as the haulage truck drivers in Uganda [6,7]. Indeed, these drivers were designated a core-risk group i.e., deemed to be more likely to introduce the disease to communities, by public health authorities, given their movement over long distances within a relatively short period [6,8]. We define a core risk group as a population that can act as a unique source of infection for the broader population [6,9,10]. Beyond introducing the infection to the general population, they can also be vulnerable to exposure and worse disease outcomes [11]. On the other hand, haulage drivers are integral to the supply chain and by extension the economy, especially in landlocked countries such as Uganda. For example, it was estimated that disrupting the haulage-based supply chain would result in a 3.3% GDP contraction for the East African region [12]. Therefore, to strike a balance between these two trade-offs, the Ugandan government implemented screening and contact tracing tailored to the haulage truck drivers [13]. However, this was not without controversy, including potential violation of individual rights[13], as it involved joint public health and security agency teams[6], often with media coverage. This heightened public anxiety and stigmatization towards this group [14]. This disproportionate scrutiny towards haulage drivers is not new. In the 1990s, these drivers were considered a core risk group for HIV spread linked to the sex trade on their routes [6,13]. What was new, and probably un-justified, was the use of strategies applied for a sexually transmitted disease (HIV) to respiratory infection (COVID-19).

To identify long-term solutions that protect the rights of such vulnerable but essential workers, we must examine the intended goals and actual outcomes of population-targeted interventions. For example, it is critical to understand if the targeted interventions only reduce the potential for introducing a pathogen, or if it also contains the potential to reduce the spread of the pathogen within the country. This distinction is well studied in HIV transmission, which is a chronic infection, but is yet to be fully explored in an acute infectious disease such as COVID-19 [15,16]. It is against this background that we examined the risk attributed to haulage drivers and the likelihood of introducing the virus and spreading it to communities in Uganda. We also examine if the interventions that were used including: a) testing for COVID-19 and waiting for results at the border points, b) testing and not waiting at the border points but receive results through digital contact tracing tools c) and targeted vaccination. It is worth noting that in Uganda, testing was tailored to specific groups, no mass testing of the population was performed. Consequently, there was never an estimate of the true prevalence.

In this regard, models can be used to characterize disease outbreaks [17] by employing mathematical formulae that address questions such as: a) Where and how did the outbreak start? b) How is it likely to spread? and c) did our interventions achieve the desired outcomes? [18] In the process we can identify population characteristics, at-risk groups [15] and their contribution to the outbreak size. By using large empirical datasets relevant to an epidemic outbreak i.e., national surveillance test results dataset, such models can be refined [17] to inform future pandemic preparedness strategies.

Here, we focus on responses targeted to haulage drivers to unravel their role in the epidemiology of COVID-19 during the Delta wave [19]. To do this, we answer the following questions: a) Does the data support the notion that haulage drivers were a core-risk group, on the account that districts linked to heavy haulage traffic registered a disproportionately higher apparent prevalence of COVID-19 cases? b) Were the targeted interventions such as i) mandatory testing and waiting for results at ports of entry or ii) mandatory testing and not waiting, effective? and c) Could alternative approaches such as digital contact tracing or targeted vaccination have yielded better results?

## Materials and Methods

### Study design

We analysed the COVID-19 national surveillance test results dataset for Uganda. Among the categories of groups tested, we focus on the haulage truck drivers who were designated a core-risk group (contracting and spreading), given their wide contact structure [6]. We hypothesised that interventions tailored to haulage, such as targeted COVID-19 screening, and manual contact tracing could limit the risk of this group spreading the virus to the public. We started by fitting models to the national COVID-19 test result dataset to identify models with the best fit. These models, among others, generate verifiable output on the incidence rates and testing rates. Then, using counterfactual concepts, we estimate outbreak parameters, such as the number of cases per location at a given time and assess whether locations linked to haulage had more cases during the Delta wave. We then simulate the impact of interventions based on the case load for specific locations/districts. Finally, we assessed the impact of an intervention or a combination of interventions in reducing this case load. Here, we assume that the movements of drivers on the road represent a contact structure with communities, and we acknowledge that testing completeness between groups may vary. All this is done using a deterministic dynamic mathematical models [20] on the national surveillance dataset for the Delta wave (10 May 2021 to September 2021) N=625,422 [19]. It is noteworthy that we focused on the Delta wave because of the good model fit to the data, and thus use the counterfactual concepts to investigate the study objectives.

### Data sources

#### COVID-19 testing dataset

We used data from the Electronic Result Dispatch System (eRDS) database hosted by the Ministry of Health (MOH) in Uganda [21]. This database contained all COVID-19 testing results received from the 22 MOH-accredited diagnostic laboratories across the country. In January 2023, the eRDS database contained 2.9 million test results covering all the pandemic waves in Uganda. All data used were anonymised, and metadata variables include age, sex, the reason for testing, occupation (risk group), testing district, district of origin, and vaccination status. The test results used here are based on the polymerase chain reaction (PCR) test [22] and we specifically focus on the Delta wave. Mandatory testing of haulage at the points of entry and exit (POE) was one of the interventions used the inland districts (Figure 1) [23].

**Figure 1.**
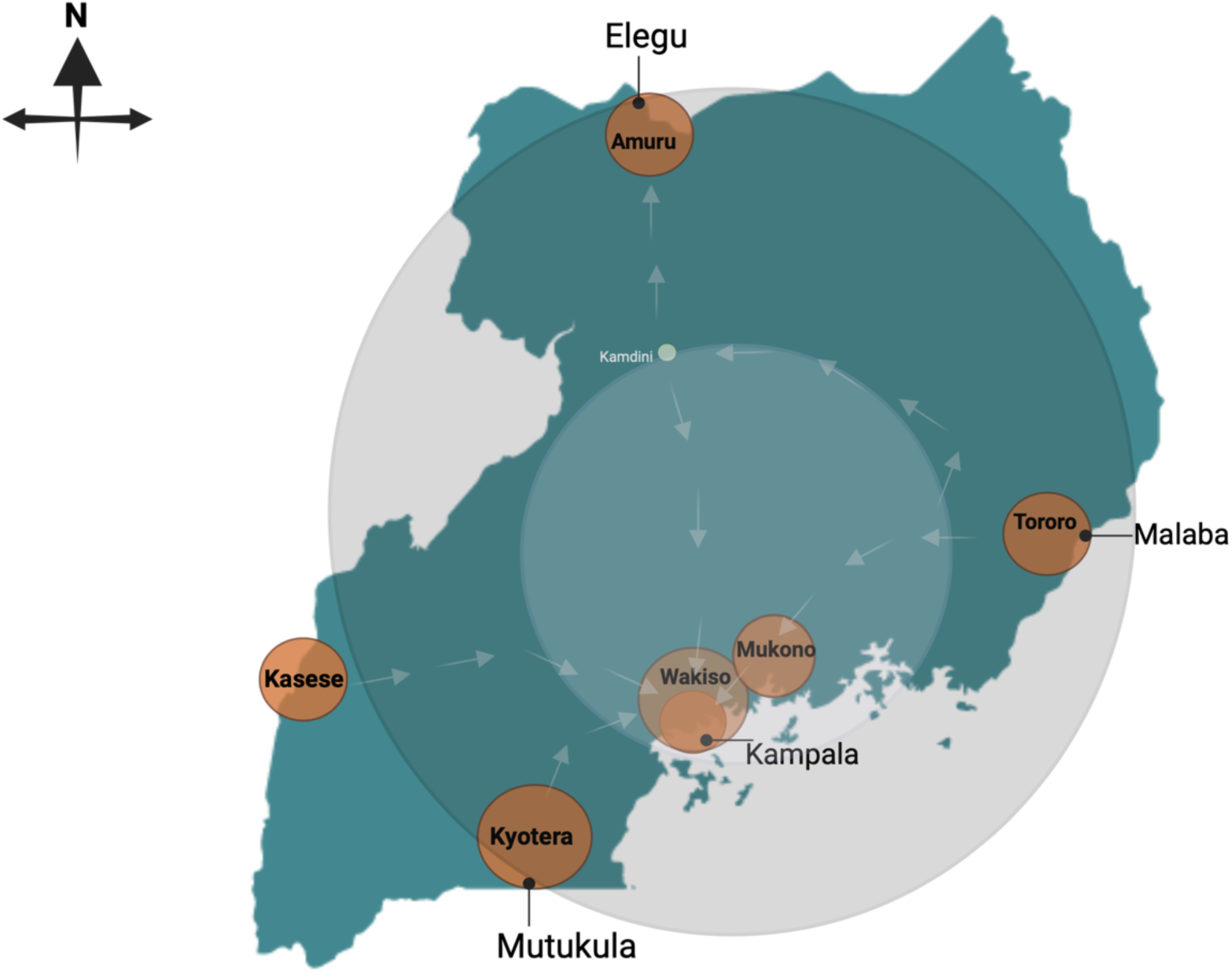
Map of Uganda showing the outer and outer ring strategy employed during the COVID-19. The outer ring included low-population density border district with points of entry and exits. The inner ring included high-population density districts protected from the risk of introduction via haulage drivers

#### Digital contact tracing data tailored to haulage

The network of haulage based on movements along the road network in Uganda was obtained from the THEA-GS project [13], which was integrated with the eRDS. THEA-GS is a digital contact tracing (DCT) tool tailored for the haulage sector [24], is mobile application collects time-stamped GPS data. The application tool uses the national COVID-19 surveillance test results in the eRDS to deliver a location-based contact tracing for the haulage sector. The THEA-GS dataset contained 62 million time-stamped GPS points linked to individual trucks, which were used to reconstruct the haulage traffic flow as proxy for contact structure between drivers and communities along the road network [24]. Haulage truck drivers on this system used six designated points of entry and exit (POE), including Malaba (border with Kenya), Elegu (border with South Sudan) and Kyotera (border with Tanzania). These are also categorised here as the districts associated with the haulage routes.

### The dynamic mathematical models

We used a population structure (see Table1) and a susceptible-infected-recovered (SIR) framework to model the characteristics of COVID-19 in six selected districts connected to the haulage sector in Uganda. The deterministic model was built on differential SIR equations, implemented using the *deSolve* package (version 1.40) in R [25] and we assume the following; a) routine testing was infrequent for most Ugandans, with only key groups such as health workers, travellers, and truck drivers eligible for regular testing, b) individuals testing positive would adhere to regulations, including self-isolation, hand hygiene, and mask usage, to limit transmission, c) the model was developed for a small-scale contact network, and d) given that long-distance travel was significantly restricted during the pandemic waves, we expect that the population in the districts remained relatively constant throughout both day and night.

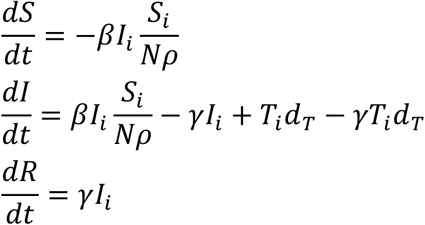

Where on day *i, S*, *I* and *R* are the number of Susceptible, Infected and Recovered individuals in the population, *N* is the district population size, 𝜌 is an adjustment factor applied to describe the proportion of the population that would present for COVID-19 testing set to 0.05, 𝛽 is the transmission rate, 𝛾 is the recovery rate = 1/5.7 [26], *T_i_* is the number of infected truck drivers on day *i* and *d_T_* is the mean duration that the truck drivers stop at a crossing point which is taken as the time spent getting tested for COVID-19 (*Intervention A*).

𝛽 is fitted from the national COVID-19 testing dataset by fitting an exponential model to the infection numbers prior-to and following a notable time point such as lockdown, so we use two values of 𝛽, one to describe the upwards part of the curve and the second the declining case numbers. We expect transmission to be linked to population density, however if we observe high transmission or high number of cases in a district with low population density, but linked to haulage, we deem haulage as a likely causal pathway. We therefore separately fitted transmission rates to three different types of areas – one covering the capital and largest city of Kampala, one for highly densely populated districts and the third for less densely populated districts (<500 people / km^2^) including districts that host ports of entry and exit.

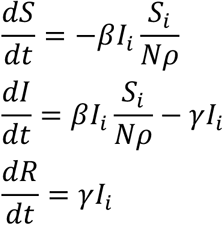

This would simulate drivers subjected to a very quick test and continuing their journey immediately to be contacted digitally (via THEA-GS) rather than waiting for results (*Intervention B*), with drivers isolating in the event of a positive result. In the third model the driver is not tested and can return to their home district and contribute to community spread.

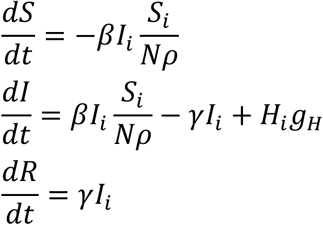

*H_i_* is the number of truck drivers returning to their home district on day *H* and *g_H_* is the proportion of infection remaining when the driver returns home (by default 0.5). We also use the scenario of no intervention i.e., not testing and no stopping at the POEs as the baseline for comparing all the intervention scenarios including vaccination *(Intervention C)*. Vaccination is implemented by assuming that the transmission rate will be reduced by a factor describing the efficacy of the vaccine ((1-ν)β).

#### Mandatory testing (Intervention A)

Haulage truck drivers were required to have a negative test certificate to travel. As such testing was primarily done at POEs in border districts of Tororo, Amuru and Kyotera, where they had to wait for the results.

#### Digital contact tracing (Intervention B)

A location-based contact tracing tool using time-stamped GPS points and test results was used to automate the contact tracing process [13,24]. Here the drivers were permitted to undergo testing and continue their journey, and thereby returning to their home district. They received their results through the THEA-GS mobile application [24], which also provided instructions regarding self-isolation. Additionally, the movement data in THEA-GS could also be used to monitor compliance with instructions to self-isolate.

#### Vaccination (Intervention C)

The public vaccination campaign commenced in April 2021 [27], and as part of this effort, truck drivers were required to be vaccinated. The vaccination certificate become a mandatory requirement for driving, in addition to presenting a negative test result.

### Model calibration

In order to generate estimate parameters from the model, we fitted a transmission model against the COVID-19 incidence as reported [28] from the eRDS between July 2020 and July 2022. Since this paper is investigating attributable risk and the impact of targeted interventions on number of infected cases, we restrict the model to 90 days within which each of the waves.

### System initialization

The initial value for *I* is the mean prevalence of infection at the point defined as the start of the epidemic wave – with one value for Uganda and a separate value for just Kampala. We do sensitivity analysis around these initial *I* value to give a counterfactual to explore the impact of truck drivers in an instance where the disease was present at high levels in neighbouring countries, but low levels in Uganda. We decrease these *I* values by percentages ranging from 0-100% of their baseline values.

### Ethical considerations

The research involving human participants underwent review and approval processes by several ethical committees, including the School of Public Health Higher Degrees, Research, and Ethics Committee at Makerere University (approval number SPH-2021-35), the Uganda National Council for Science and Technology (approval number HS156ES), and the Human Ethics and Research Committee at the Easter Bush Campus (HERC_538). The approvals included access to the national testing data as part of the THEA project. Written informed consent sought digitally was obtained from all participants prior to their inclusion in the study.

## Results

### Descriptive summary of dataset

Of the 2.9 million COVID-19 test results in the eRDS, 40.2% were female and 58.6% were male and the rest were undefined genders. The age range was 1-90 years and a median age of 35 for males and 28 for females. 6.3% of test results were positive, with the median age group for positive cases being 30-40 years. Non-Ugandan travellers accounted for 8.6% of the test results. Among the tested drivers, 2.1% were positive compared to health workers at 16.6% (Table 2). Overall, haulage truck drivers accounted for 0.036% of all positive tests in the eRDS database. Here, focusing on the Delta wave, as shown in Figure 2, a high proportion of tests and cases were registered in the eRDS. It is important to note that data hygiene was an issue, especially a lack of harmonized format for entries, which resulted in numerous variants of categories.

**Figure 2.**
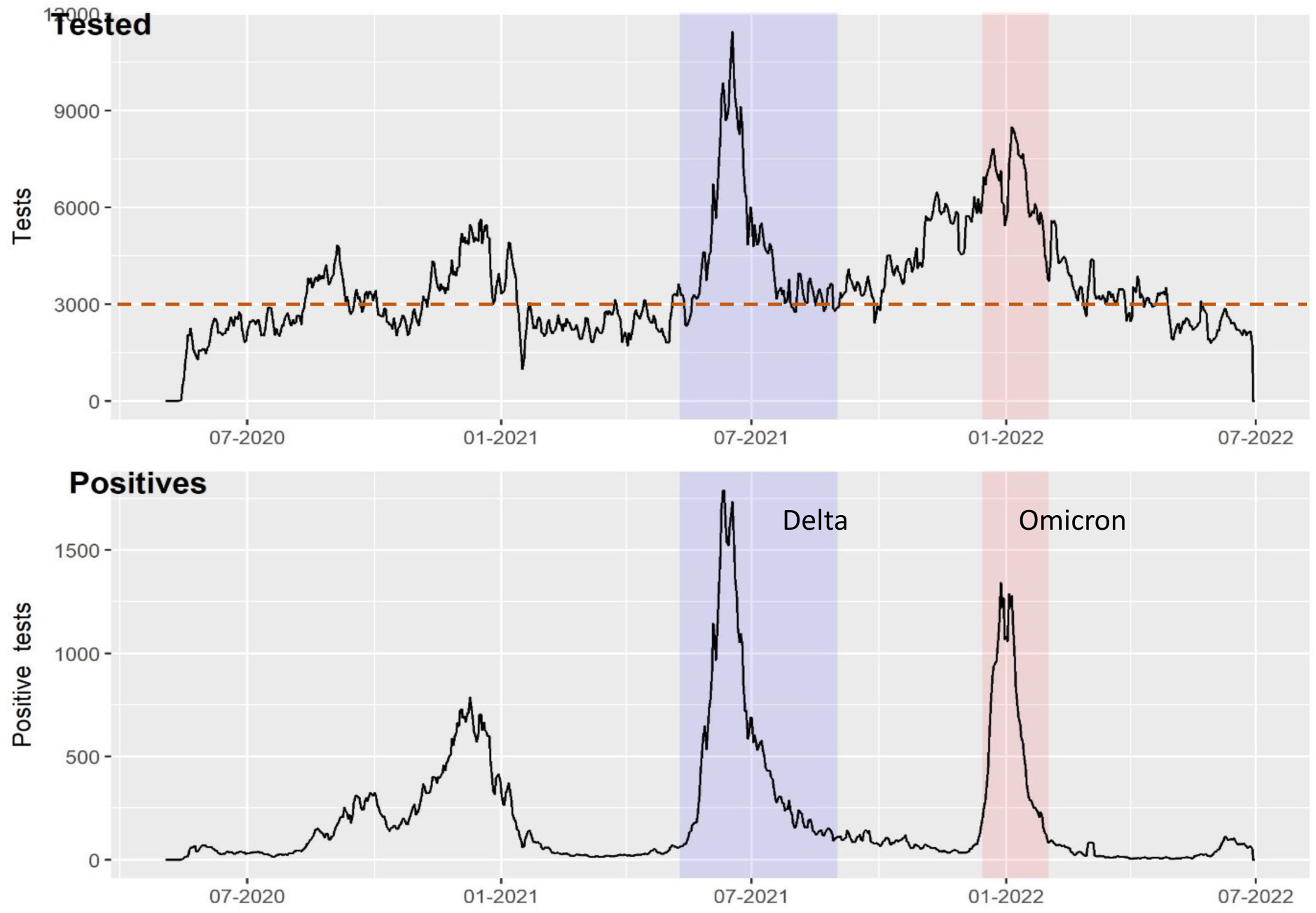
A line graph showing the total numbers tested and positive tests during the Delta and Omicron waves of the pandemic. The dotted line shows the average testing rate in the study period. The plot was generated with ggplot in R version 4.2.2.

**Table 1.** Parameter estimates used to calibrate the model.

| Parameters/variables | Values | Comments |
| --- | --- | --- |
| Baseline transmission rate | Fitted |  |
| Susceptible population |  |  |
| Kampala | 1,680,600 | https://www.citypopulation.de/en/uganda/admin/ |
| Wakiso | 2,915,200 |  |
| Mukono | 701,400 |  |
| Tororo | 597,500 |  |
| Amuru | 216,800 |  |
| Kyotera | 261,000 |  |
| Testing rate | 3000/day | ~ 108 drivers tested per day although not explicitly used for modelling |
| Initial value of $I$: | | |
| Kampala | 17 | Fitted based on the infection rates in areas of different population densities |
| Wakiso | 2.85 |  |
| Mukono | 0.685 |  |
| Tororo | 0.583 |  |
| Amuru | 0.291 |  |
| Kyotera | 0.351 |  |
| Population adjusted factor ($\rho$) | 0.05 | |
| Generation time ($\Gamma$) | 5.7 | Salzburger *et al.* [26] |
| Recovery rate ($\gamma$) | $1/\Gamma$ | Generation time form |
| Transmission rate ($\beta$) | Fitted | |
| Vaccine efficacy | 0.75 | The proportion of vaccinated individuals that are protected from infection |

**Table 2.**
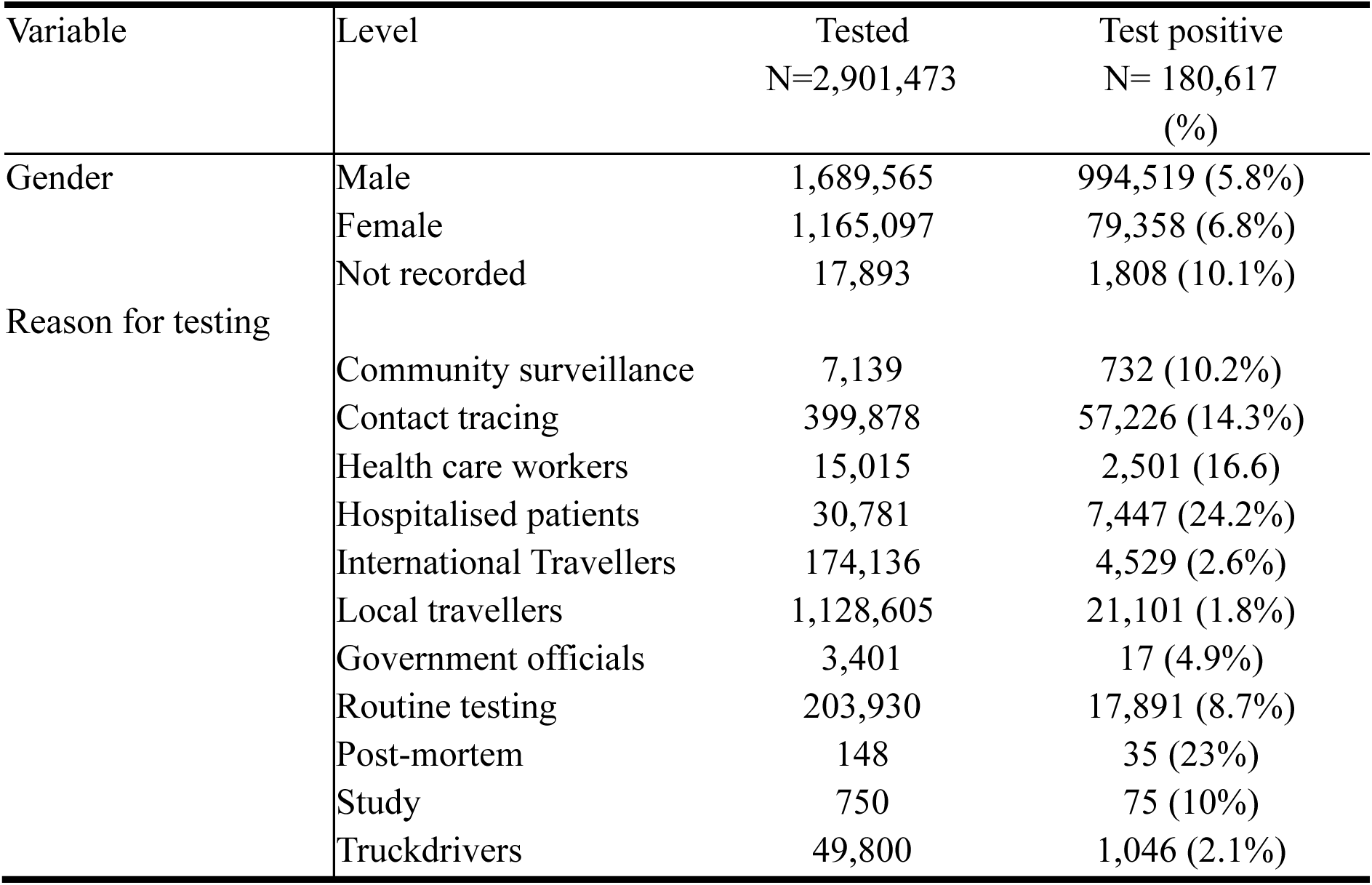
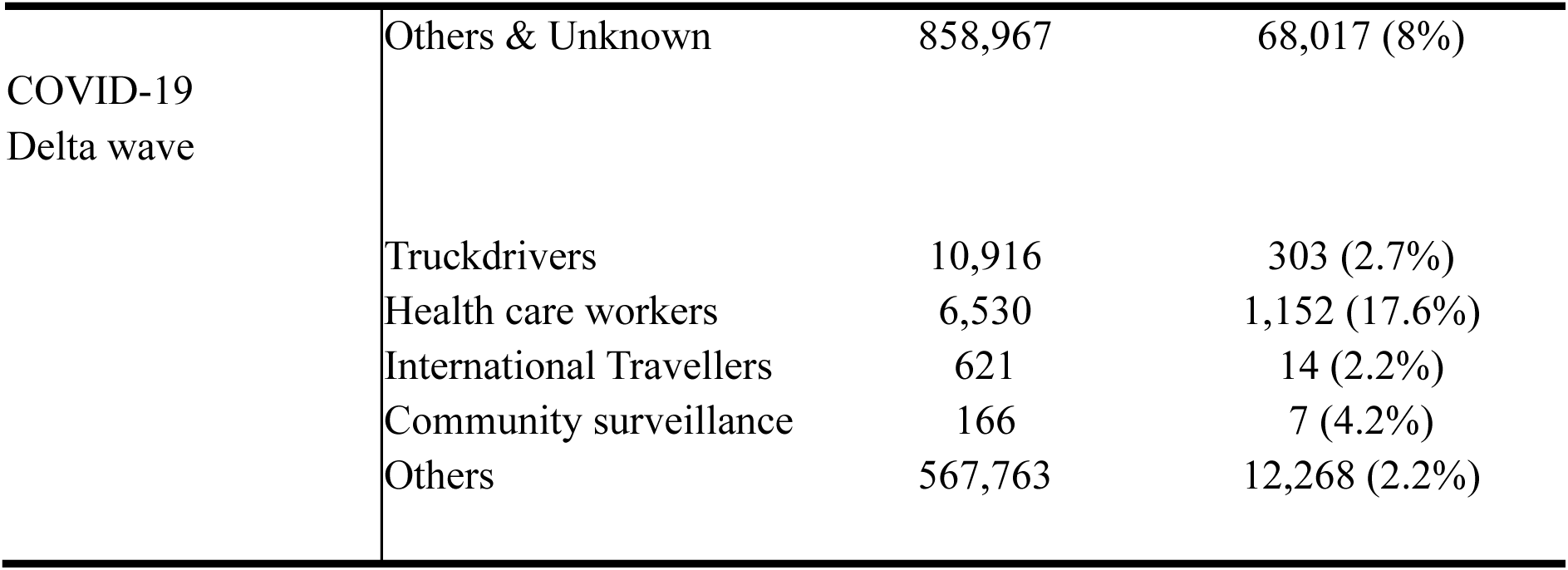
SARS-CoV-2 Testing rates and reasons for testing.

### Haulage-associated epidemic characteristics

To assess the case load for districts and regions linked to haulage, we assess epidemic characteristics of based on population density. Here the fitted models show a comparable (*R_0_*) between high and low. The findings show that interventions implemented during the Delta wave effectively reduced the reproduction number (*R_0_*) for the whole country from 1.98 pre-lockdown to 0.78 post-lockdown (**Table 3 and Figure 2**). The trend was similar for the low-density border districts and the medium-density peri-urban districts but it is worth noting that the (*R_0_*) in peri-urban districts of Mukono and Wakiso was higher pre-lockdown than low density districts like Amuru, Kyotera and Tororo.

**Figure 2a.**
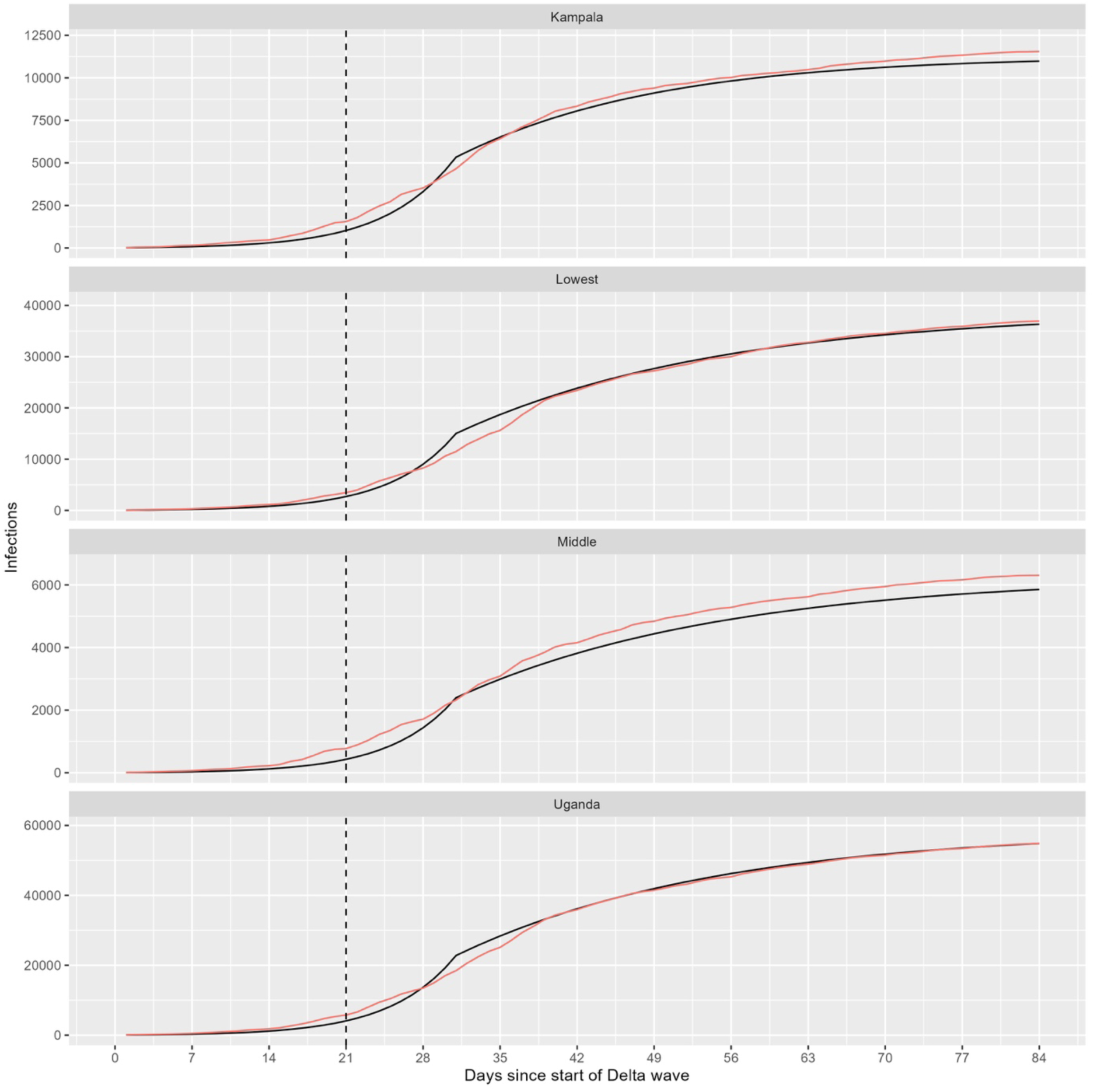
Results of fitting the Delta wave to Uganda, Kampala, and mid (peri-urban) to low (border districts) population density district. The black and pink lines represent the empirical data and model respectively and highlights the similar fit in each population group. The dotted black line represents the day when the lockdown was instituted.

**Table 3.** Fitted R0 values to the different study regions.

| Location | $R_0$ (95% CI) | |
| --- | --- | --- |
| Region | **Rising phase – pre-lockdown**<br>(10-5-2021 - 08-06-2021) | **Control phase – post-lockdown**<br>(09-06-2021 – 30-06-2021) |
| Uganda | 1.98 (1.92 – 2.04) | 0.776 (0.745 – 0.807) |
| Kampala | 1.96 (1.89 – 2.03) | 0.816 (0.788 – 0.843) |
| Low population density | 1.98 (1.92 – 2.04) | 0.756 (0.723 – 0.791) |
| Mid population density | 2.01 (1.93 – 2.09) | 0.800 (0.774 – 0.825) |

**Table 4.**
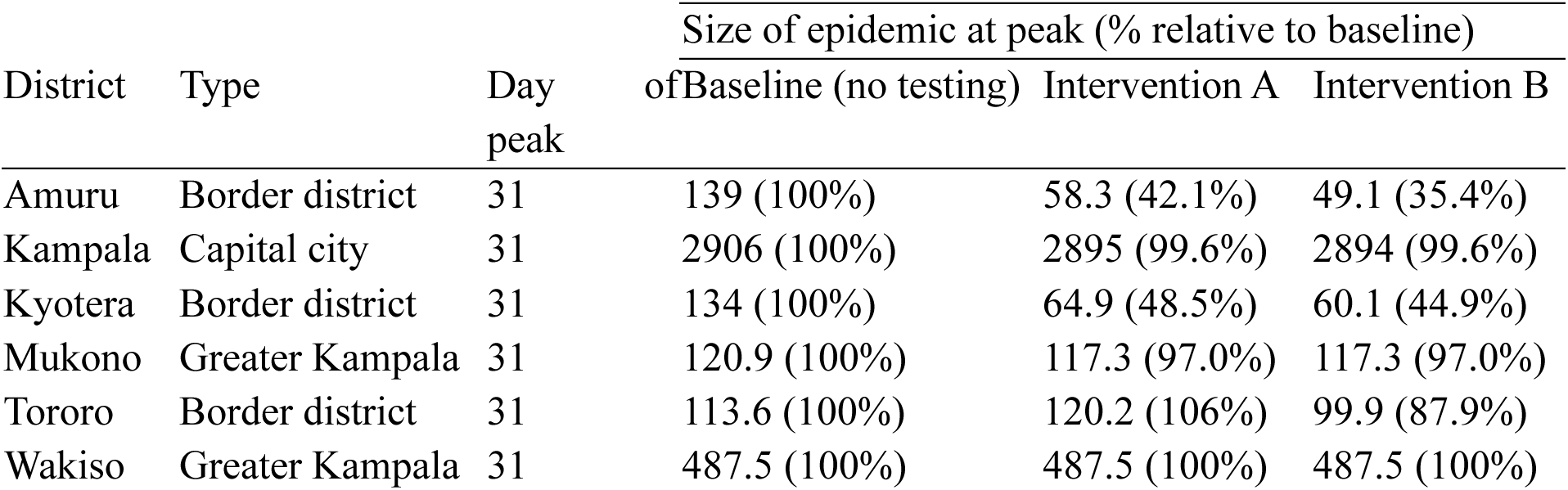
Model output on the effect on the size of the epidemic among haulage drivers at its peak for a given intervention during the Delta wave.

| District | Type | Day peak | Size of epidemic at peak (% relative to baseline) |  |  |
| --- | --- | --- | --- | --- | --- |
|  |  |  | of Baseline (no testing) | Intervention A | Intervention B |
| Amuru | Border district | 31 | 139 (100%) | 58.3 (42.1%) | 49.1 (35.4%) |
| Kampala | Capital city | 31 | 2906 (100%) | 2895 (99.6%) | 2894 (99.6%) |
| Kyotera | Border district | 31 | 134 (100%) | 64.9 (48.5%) | 60.1 (44.9%) |
| Mukono | Greater Kampala | 31 | 120.9 (100%) | 117.3 (97.0%) | 117.3 (97.0%) |
| Tororo | Border district | 31 | 113.6 (100%) | 120.2 (106%) | 99.9 (87.9%) |
| Wakiso | Greater Kampala | 31 | 487.5 (100%) | 487.5 (100%) | 487.5 (100%) |

### Assessing haulage targeted intervention strategies

#### Border districts associated with haulage

The model shows a higher epidemic peak for the baseline i.e. no testing and therefore no stopping at the POEs of the border districts. Implementing Intervention A, i.e. truck drivers tested at POEs and waiting for a result (blue line in Figure 3), there was a reduction in cases, with the most significant reduction observed in districts with POEs like Kyotera (41%) and Amuru (48%) (Table 2). However, in Tororo district, Intervention A resulted in a case load higher than the baseline (Figure 3). Intervention B where testing is done, and results delivered via a DCT mobile application showed the largest reduction in cases within border districts. For example, Amuru (50.9%) and Kyotera (39.9%). Here too the characteristics of Tororo district are different from other border districts, showing only an 11% reduction in cases relative to the baseline intervention. Interestingly, Intervention C which combines vaccination with either baseline, intervention A or B appears to have the largest effect on the number of infections in Tororo district (Figure 3).

**Figure 3.**
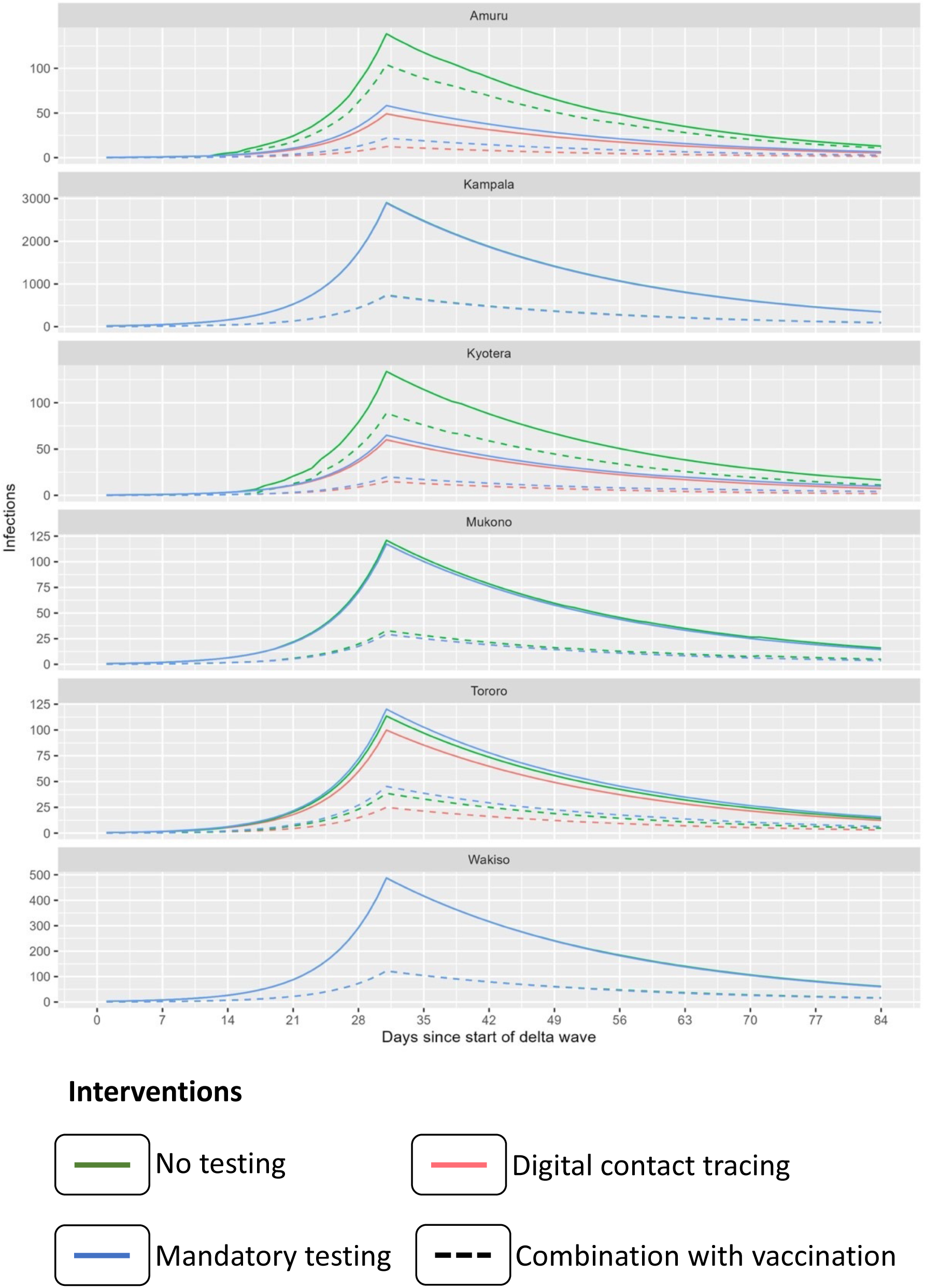
During the Delta wave, we evaluated the impact of three strategies across six modelled districts: Blue is Intervention A: Truck drivers were tested at the POE or designated seclusion points and were required to wait for their results at that location. They could only proceed if their test result was negative. Red is Intervention B: in this scenario, a digital contact tracing tool was utilized. Truck drivers were automatically contacted through a mobile application to relay a) test results, b) public health guidance, and c) geofencing to evaluate compliance. Green (No Testing - No Intervention): This scenario represents no testing, and hence, the health status is unknown. All individuals are allowed to travel freely. This serves as the reference or control for our analysis. The dotted lines represent a combination of each respective intervention with targeted vaccination using a vaccine that is 75% effective (Intervention C).

#### Peri-urban districts

At the peak of infection, 31 days since the start of the Delta wave, we note a negligible difference between Intervention A and the baseline especially in Mukono district. Although intervention C resulted in a significant difference in the number of infections in both Mukono and Wakiso, in Mukono the difference between baseline and Intervention A remains negligible. It is worth noting that in urban settings in Kampala and Wakiso, truck drivers were not allowed to travel without a test, hence this intervention is not factored in.

#### Kampala district, the capital city

Similarly, in the capital city, there is a negligible difference between Intervention A and B relative to the baseline. The profile of change in infections when Intervention C was implemented was similar to Wakiso although the actual case load in Kampala was much higher (**Figure 3**).

Overall, if vaccination had been implemented as a standalone intervention, it would have resulted in a 1.4% reduction of cases in the districts liked to haulage.

### Using the haulage movement network: potential additional risk form inland movements

Figure 4 illustrates the impact of the onward movements of trucks from border crossing points. The baseline analysis focused on the primary increased risk from truck drivers, which shows that this impact was concentrated within POE districts. Although some in-land districts show some accumulation of risk following the incorporation of the truck network and the additional transmission resulting, it is far lower than it is in the border districts (Figure 4). Furthermore, the impact of varying the initial (*I*), simulating a lower rate of introduction is assessed to show the changes in truck driver-attributable infections. The result suggests that the impact of truck drivers on infection size is at its maximum (75%) when (*I*) is at its lowest (0.03), as would be the case at the beginning of the Delta wave. This means that if the disease had been contained and kept out then truck drivers would have had the biggest impact on potential transmission. This impact is notably pronounced in Tororo and Amuru districts and, to a lesser extent, in Kyotera (Figure 4 C&D). This finding suggests that targeted intervention is effective in averting a substantial number of new infections when implemented during the early stages of an outbreak. Figure 4B suggests the optimal level to intervene appears to be when the prevalence (initial value) is 3% and 9% beyond which if no intervention is implemented would result in a 50% and 26% increase in cases.

**Figure 4.**
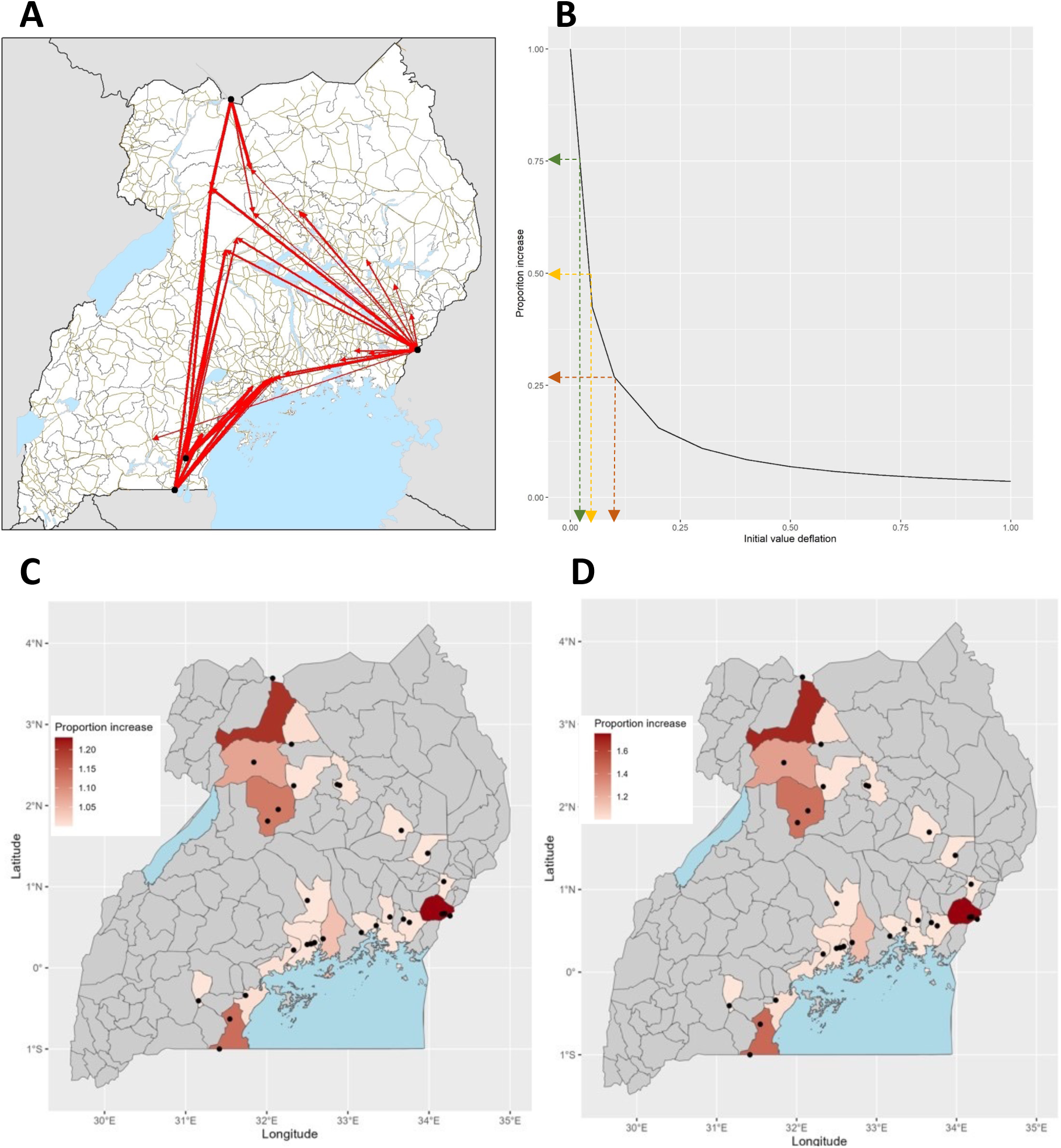
A The haulage truck network was generated using time-stamped GPS data from the digital contact trace system THEA-GS [24]. Here we show the links for the most used routes, and the ability of truck drivers to move disease inland. In addition to these observed links, we put in a link from Amuru to the one node without a vertex but made this a low-frequency vertex. B shows how the impact of interventions is dependent on the level of infection in the population. For example, if the intervention (deflation) on the x-axis is done at 0.03, it is possible to achieve a 75% increase in the proportion of cases averted. C & D shows the proportion increase in caseload attributable to onward movement of the haulage drivers during the Delta wave

## Discussion

In this study, we sought to examine the role of haulage truck drivers as a core risk group in the epidemiology of the Delta wave of COVID-19 in Uganda. By doing, we understand how intervention can be optimally deployed to reduce the potential for introduction and spread to the rest of the populations with limited infringement on the individual rights of truck drivers in Uganda. Therefore, the overarching aim was to learn from the responses to inform current and future pandemic preparedness strategies in similar settings. This is important because the haulage sector is integral to national and international supply chains and by extension, their economies [12]. The occupational and public health risks associated with this group are inherently linked to the breadth of their contact structure and could result in excessive public scrutiny during disease outbreaks with welfare consequences. Although our findings support the notion that haulage truck drivers were a core-risk group [6] during the Delta wave, our findings suggest that this risk was limited to specific spatial and temporal contexts.

### Haulage as a core risk group

Here, we apply the core risk categorization to haulage truck drivers because although this group accounted for only 0.036% of all cases in the eRDS, border districts such as Tororo, which are uniquely associated with heavy haulage traffic, experienced a 12.02% increase in incidence during the Delta wave and the least change in case load following interventions. This suggests that truck drivers were a risk group and some targeted interventions may have exacerbated their risk [29]. Indeed, our analysis of haulage-targeted interventions shows a pronounced impact on incidence in border districts. For instance, while Intervention A, which involved testing and waiting for results at the border, resulted in a ∼50% reduction of cases in Amuru and Kyotera, it resulted in a 6% increase of cases in Tororo district relative to the baseline with no intervention. This difference in outcomes likely reflects the variation in the number of truck drivers using the border crossing and the congregation that resulted while waiting for results. Tororo, with large volumes of trucks from Kenya destined for South Sudan and the Democratic Republic of Congo, represented a greater risk of introduction to the general population of Tororo. By this measure, truck drivers appear to have been a core risk group [6]. However, the risk and impact were generally localized to border districts and transient, with the highest impact at the onset of the outbreak. Future targeted interventions could benefit from such context to inform public health decision-making and responses that limit generalised stigmatization of the haulage sector [6]. In this regard, consideration of factors that inherently render haulage drivers vulnerable to exposure is crucial. For instance, our data indicates that a typical driver is male in his 40s or 50s [30]. Their occupation requires long periods spent sitting alone in truck cabins, often with suboptimal dietary habits [11]. Furthermore, they are frequently exposed to chemical and biological pollutants, coupled with stressors from road infrastructure [11]. Such a lifestyle can contribute to the early onset of non-communicable diseases, including mental health issues, diabetes, heart disease, and kidney disease [11]. When these conditions intersect with infectious disease pathogens, it becomes evident why this group can easily be classified as a core-risk group [31]. It is therefore imperative to update occupational health policies to promote awareness, early diagnosis, and contextualized communication of interventions. Such strategies should be integral to current preparedness strategies to support public health and the economy while safeguarding the rights of drivers in haulage sector.

### Effectiveness of targeted interventions

The objective of an intervention to an infectious disease outbreak is to minimise the probability of an introduction and spread of a pathogen [32]. In Uganda, the mass testing program began in April 2020, allowing haulage truck drivers to travel only if they tested negative for COVID-19. Therefore, testing facilities targeting haulage were established at POEs in border districts [13]. Our research findings show that while this intervention reduced cases in Kyotera and Amuru, it resulted in an increase in Tororo district. This rise reflects the effect of unintended delays, with drivers waiting 24-36 hours for test results at the border, disrupting the haulage chain [14]. At its worst, the queue of trucks was 47 Km at the Kenya-Uganda border in Malaba [14], leading to overcrowding and likely increasing the transmission. When haulage drivers were allowed to test and proceed with their journey and received their results via a contact tracing mobile application (Intervention B), the case load in Tororo district was reduced by 11% relative to the baseline without intervention. The reduction was even much higher in Amuru and Kyotera. The variation in case load between the three border districts following targeted interventions suggests haulage played a role in the epidemiology, albeit to varying degrees across districts. The variance in caseload between Tororo and Amuru districts might also be attributed to shorter transit times at Elegu port, since trucks and drivers [24] will have undergone pre-inspection and clearance at Malaba port, respectively

According to the European Centre of Disease Prevention and Control [33] the primary objective of the vaccination campaign was to reduce the pressure on health care systems and facilitate the re-opening of society. Similarly, in Uganda where the vaccination campaign began in March 2021 [12,34], the vaccinated population primarily received AstraZeneca (93%), 5% Moderna and 2% Pfizer [12,34]. At this point, truck drivers were required to have a valid vaccination certificate along with a negative test to travel. Our findings indicate that vaccination alone would have reduced the overall case load by 1.4%. This is probably because the vaccines were primarily aimed at improving case outcomes [33], easing pressure on the healthcare system [29], not preventing transmission. This does not suggest that vaccination as an intervention was ineffective; rather our findings suggest that better outcomes with case numbers could be achieved with proper timing and combination with other interventions as discussed in the following sections.

### The potential for alternative intervention

Digital contact tracing (DCT) has emerged as a potential epidemiological tool to support reopening of economies [35] by limiting transmission while maintaining functional supply chains [36]. Our result indicates that deploying a digital contact tracing tool [24] could significantly reduce the caseload by 35% in border districts like Amuru and by 44% in Kyotera. This would allow drivers to continue with their journey and access results through mobile phone applications, thereby reducing border transit time, overcrowding and subsequent transmission. Similarly, we did not observe a much lower reduction (16.9%) of cases in Tororo district, this too likely reflects the role of the volume of traffic in the effectiveness of interventions [13]. One the other hand we observed that this intervention was not effective in decreasing the caseload for densely populated urban areas such as Kampala, Mukono, and Wakiso in the central region of the country. This is probably due to the presence of multiple risk groups, including international travellers and health workers. Thus, it can be argued that haulage-targeted digital contact tracing interventions may primarily benefit border communities.

Some studies have indicated that DCT may be less effective in urban settings, primarily due to low resolution from potential barriers such as building structures affecting GPS or Bluetooth signals [37]. The reasons for the limited utility observed in this study remain unknown and could be a potential area for further investigation.

Our results indicate that a further reduction in the case load can be achieved by combining all the above interventions with vaccination. For example, between 60% and 80% reduction in cases could have been achieved in Tororo and Kampala respectively. Similar results would be achievable in Amuru and Kyotera districts. Studies conducted elsewhere have shown the benefit of combining vaccination [38]. In South Africa, they have shown that such an approach offers better outcomes including potentially suppressing an outbreak [38]. Beyond combining interventions, the timing of an intervention is critical [28]. Here, we show that the risk associated with truck drivers is transient and highest at the beginning of an outbreak when community prevalence is at its lowest. Therefore, implementing a combined intervention when the prevalence is at 3% could have resulted in a 75% reduction in case load.

#### Study limitations

a. Our modelling approach did not consider the onward impact of this group on all other districts in Uganda. This constraint limits the generalizability of our findings beyond the specific districts examined in the study. It is possible that each district has unique contact structures, leading to different and unaccounted dynamics.
b. Our analysis primarily centres on the Delta wave of the pandemic in Uganda. While we also investigated the Omicron wave, the model’s accuracy was not robust enough to facilitate subsequent counterfactual analysis. With these caveats, our finding suggests that the risk introduction associated with haulage during this wave was limited to border districts, even then, interventions contributed to it. We also find limited evidence that haulage drivers played a role in onward transmission within the central regions

### Relevance to future pandemic preparedness strategies

Haulage truck drivers are at a unique intersection between public health-economic interests, and historic public biases. During the pandemic such interests diverge, and biases amplified, leading to unwarranted scrutiny of this group of essential workers. These pressures and biases add to the multitude of challenges and health vulnerabilities that truck drivers already face [11]. Therefore, our findings contribute to the evidence necessary for updating policies and strategies aimed at building resilient logistics supply chains capable of supporting populations during pandemics. In this study we have characterized the risk and show that it was place and time-specific, in some cases amplified by the very interventions put in place. Therefore, the implementation of targeted and early interventions is crucial. For instance, despite Tororo, Kyotera, and Amuru being border districts associated with haulage traffic, they each require context-specific interventions to effectively manage the risk. Ultimately all this contributes to the enhancement of occupational health within the haulage sector by highlighting gaps.

## Conclusion

Our findings support the notion that haulage truck drivers were a core-risk group during the Delta wave, but this risk was transient and likely exacerbated by the interventions, such as testing and waiting for results at points of entry (POE) in districts like Tororo. Therefore, high-volume border crossings require tailored interventions. In this regard, a combination of interventions, such as digital contact tracing and vaccination, could have served as more effective alternatives in the public health institution’s response toolkit.

## Data Availability

The dataset use is available upon request from the Ministry of Health of Uganda

